# Modeling and Forecasting of Covid-19 growth curve in India

**DOI:** 10.1101/2020.05.20.20107540

**Authors:** Vikas Kumar Sharma, Unnati Nigam

## Abstract

In this article, we analyze the growth pattern of Covid-19 pandemic in India from March 4^th^ to July 11^th^ using regression analysis (exponential and polynomial), auto-regressive integrated moving averages (ARIMA) model as well as exponential smoothing and Holt-Winters smoothing models. We found that the growth of Covid-19 cases follows a power regime of (t^2^,t,..) after the exponential growth. We found the optimal change points from where the Covid-19 cases shift their course of growth from exponential to quadratic and then from quadratic to linear. After that, we saw a sudden spike in the course of the spread of Covid-19 and the growth moved from linear to quadratic and then to quartic, which is alarming. We have also found the best fitted regression models using the various criteria such as significant p-values, coefficients of determination and ANOVA etc. Further, we search the best fitting ARIMA model for the data using the AIC (Akaike Information Criterion) and provide the forecast of Covid-19 cases for future days. It was noticed that the ARIMA model fits better the Covid-19 cases for small regions. ARIMA (5, 2, 5) and ARIMA (5, 2, 3) are the best possible models for modeling Covid-19 cases for March 4^th^ to July 10^th^ and June 1^th^ to July 10^th^, respectively.

## 1. Introduction

The Covid-19 pandemic has created a lot of havoc in the world. It is caused by a virus called SARS-CoV-2, which comes from the family of coronaviruses and is believed to be originated from the unhygienic wet seafood market in Wuhan, China but it has now infected around 215 countries of the world. With more than 13.2 million people affected around the world and more 575,000 deaths (As of July 14^th^, 2020), it has forced people to stay in their homes and has caused huge devastation in the world economy. (Singh & Jadaun [1], [2], Gupta et al. [3]).

In India, the first case of Covid-19 was reported on 30^th^ January, which is linked to the Wuhan city of China (as the patient has travel history to the city). On 4^th^ March, India saw a sudden hike in the number of cases and since then, the numbers are increasing day by day. As of 14^th^ July, India has more than 908,000 cases with more than 23,000 deaths and is world’s 3^rd^ most infected country. ([4]).

Since the outbreak of the pandemic, scientists across the world have been indulged in the studies regarding the spread of the virus. Lin et al. [5] suggested the use of the SEIR (Susceptible-Exposed-Infectious-Removed) model for the spread in China and studied the importance of government-implemented restrictions on containing the infection. As the disease grew further, Ivorra et al. [6] suggested a θ-SEIHRD model that took into account various special features of the disease. It also included asymptomatic cases into account (around 51%) in order to forecast the total cases in China (around 168500). Giordano et al. [7] also suggested an extended SIR model called SIDHARTHE model for cases in Italy which was more customized for Covid-19 in order to effectively model the course of the pandemic to help plan a better control strategy.

Petropoulos and Makridakis [8] suggested the use of exponential smoothing method to model the trend of the virus, globally. Kumar et al. [9] gave a review on the various aspects of modern technology used to fight against COVID-19 crisis.

Apart from the epidemiological models, various data-oriented models were also suggested in order to model the cases and predict future cases for various disease outbreaks from time-to-time. Various time-series models were also suggested in order to model the cases and predict future cases. ARIMA and Seasonal ARIMA models are widely used by researchers in order to model and predict the cases of various outbreaks. In 2005, Earnest et al. [10] conducted a research to model and predict the cases of SARS in Singapore and predict the hospital supplies needed using this model. Gaudart et al. [11] modelled malaria incidence in the Savannah area of Mali using ARIMA. Zhang [12] compared Seasonal ARIMA model with three other time series models to compare Typhoid fever incidence in China. Polwiang [13] also used this model to determine the time-series pattern of Dengue fever in Bangkok.

For Covid-19 as well, various researchers tried to model the cases through ARIMA. Ceylan [14] suggested the use of Auto-Regressive Integrated Moving Average (ARIMA) model to develop and predict the epidemiological trend of Covid-19 for better allocation of resources and proper containment of the virus in Italy, Spain and France. Chintalapudi et al. [15] suggested its use for predicting the number of cases and deaths post 60-days lockdown in Italy. Fanelli and Piazza [16] analyzed the dynamics of Covid-19 in China, Italy and France using iterative time-lag maps. It further used SIRD model to model and predict the cases and deaths in these countries. Zhang et al. [17] developed a segmented poisson model to analyze the daily new cases of six countries in order to find a peak point in the cases.

Since the spread of the virus started to grow in India, various measures were taken by the Indian Government in order to contain it. A nationwide lockdown was announced on March 25^th^ to April 14^th^, which was later extended to May 3^rd^. The whole country was divided into containment zones (where large number of cases were observed from a relatively smaller region), red zones (districts where risk of transmission was high and had higher doubling rates), green zones (districts with no confirmed case from last 21 days) and orange zones (which didn’t fall into the above three zones). After the further extension of the lockdown till May 17^th^, various economic activities were allowed to start (with high surveillance) in areas of less transmission. Further, the lockdown was extended to May 31^st^ and some more economic activities have been allowed as per the transmission rates, which are the rates at which infectious cases cause new cases in the population, i.e. the rate of spread of the disease. This was further extended to June 8^th^, with very less rules and specially the states were given the responsibility of setting the lockdown rules. The air and rail transport became open for general public. Post June 8th, we see that the restrictions are nominal with even shopping malls and religious places open for general public. Now, the responsibility of imposing restrictions lies with the respective state governments.

On the other hand, Indian scientists and researchers are also working on addressing the issues arising from the pandemic, including production of PPE kits and tests kits as well as studying the behaviour of spread of the disease and other aspects of management. Various mathematical and statistical methods have been used for predicting the possible spread of Covid-19. The classical epidemiological models (SIR, SEIR, SIQR etc.) suggested the increasing trend of the virus and predicted the peaks of the pandemic. Early researches showed the pandemic to reach its peak by mid-May. They also showed that the basic reproduction number (R_0_) and the doubling rates are lower in India, with comparison to European nations and USA. A tree-based model was proposed by Arti and Bhatnagar [18] and Bhatnagar [19] in order to study and predict the trends. They suggest that lockdown and social-distancing in India has played a significant role to control the infection rates. But now, as the lockdown restrictions are minimal, the cases in India are growing at an alarming high rate. Chatterjee et al. [20] suggests growth of the pandemic through power law and its saturation at the later stages. Due to the complexities in the epidemic models of Covid-19, various researchers have been focusing on the data in order to forecast the future cases. Chatterjee et al. [20, 21] and Ziff & Ziff [22] suggest that after exponential growth, the total count follows a power regime of t^3^, t^2^, t and 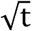 before flattening out, where ‘t’ refers to time. It can therefore be realized that there is an urgent need to model and forecast the growth of Covid-19 in India as the virus is in the growing stage here.

In India, the most affected states are Maharashtra with over 2,60,000 cases (as of 14^th^ July, 2020), Tamil Nadu (around 1,42,000 cases), Delhi (around 1,13,000 cases) and Gujarat (around 42,000 cases). The greatest number of cases per million have been seen in the national capital of Delhi (5740 cases per million). (Refer [23] for population estimates). Many states and union territories like, Kerala, Karnataka, Andaman and Nicobar Islands, Daman and Diu etc. which had recovered from majority of the cases have experienced a second wave of infections. This might be attributed to decreased travel restrictions and minimal lockdown measures. In their research, Singh & Jadaun [1] studied the significance of lockdown in India and suggested that the new Covid-19 cases would stop by the end of August in India with around 350,000 total cases. While some states may see an early stopping of new cases such as Telangana (mid-June), Uttar Pradesh and West Bengal (July-end) etc., the badly affected states of Maharashtra, Tamil Nadu and Gujarat will achieve this by August-end.

Since a proven vaccine and medication is yet to be developed by the researchers then in such a scenario, modeling the present situation and forecasting the future outcome becomes crucially important in order to utilize our resources in the most optimal way. Therefore, the article aims to study the growth curve of Covid-19 cases in India and forecast its future course. Since the disease is still in its growing age and very dynamic in nature, no model can guarantee for perfect validity for future. We therefore, need to develop the understanding of the present situation of the pandemic.

In this article, we first study the growth curve using regression methods (exponential, linear and polynomial etc.) and propose an optimal model for fitting the cases till July 11^th^. Further, we propose the use of time-series models for forecasting the future observations on Covid-19 cases. Here, we reach the best-fitted ARIMA model for forecasting the Covid-19 cases. We also compare these results with Exponential Smoothing (Holt-Winters) model. This study will help us to understand the course of spread of SARS-CoV-2 in India better and help the government and the people to optimally use the resources available to them. As this study presents the evaluation of the pandemic, it may also help the practitioners and researchers to build further models/analysis with other co-factors.

## 2. Statistical Methodologies

In this section, we briefly present the statistical techniques used for analyzing the Covid-19 cases in India. Here, we used usual regression (exponential, polynomial), times series (ARIMA) and exponential smoothing models.

### 2.1 Exponential-Polynomial Regression

Regression is a statistical technique that attempts to estimate the strength and nature of relationship between a dependent variable and a series of independent variables. Regression analyses may be linear and non-linear. A regression is called linear when it is linear in parameters e.g. *y* = *β*_0_ + *β*_1_*t*+ ∈and *y* = *β*_0_ + *β* _1_*t*+ *β* _2_*t*^2^ + *β* _3_*t*^3^+∈, ∈∼ *N*(0, α^2^), where *y* is response variable, *t* denotes the indepenet variable, *β*_0_ is the intercept and other βs are known as slopes.

A non-linear regression is a regression when it is non-linear in its parameters e.g.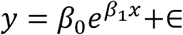. In the beginning of the spread of a disease, we see that the new cases are directly proportional to the existing infected cases and may be represented by 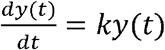, where k is the proportionality constant. Solving this differential equation, we get that, at the beginning of a pandemic,

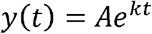

Thus, at the beginning of a disease, the growth curve of the cases grows exponentially.

As the disease spreads in a region, governments start to take action and people start becoming conscious about the disease. Thus, after some time, the disease starts to follow a polynomial growth rather than continuing to grow exponentially.

In order to fit an exponential regression to our data, we linearize the equation by taking the natural logarithm of the equation and convert it to a linear regression in first order.

We estimate the parameters of a linear regression of order *p* as following-

Let the model of linear regression of order 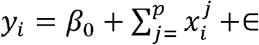 with ∈∼ *N*(0, σ^2^)and *i* = 1,2,..,*N*. Let 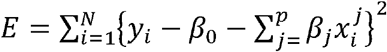 represent the residual sum of square (RSS).

We get the best estimates of these coefficients using the ordinary least squares (OLS) method. We will use this technique of the OLS in order to estimate the coefficients of our proposed model. (Refer Montgomery et al. [24])

Since we know that the growth curve of the disease changes after some time point, exponential to polynomial, we propose to use the following joint regression model with change point *µ*,

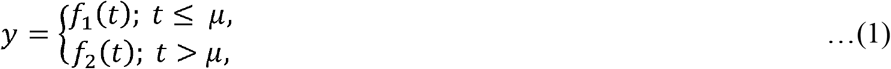

where we take 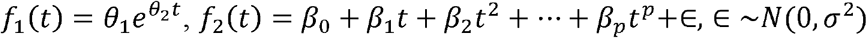 and *p* is the order of the polynomial regression model and *t* stands for the time (an independent variable).

During the analysis, we found that a suitable choice of *f*_2_ (*t*) is a quadratic or a cubic model. Once the order of the polynomial is kept fixed, an optimum value of the change point can be obtained by minimizing the residuals/errors. We can obtain the OLS estimates of the parameters of the model (1) as given below.

The least square estimates (LSEs) of the parameters, Θ = {*θ* _1_, *θ* _2_,*µ, β* _0_, *β* _1_, *β* _2_, *β* _3_,……, *β*_*p*_}can be obtained by minimizing the residual sum of squares (RSS) as given by-

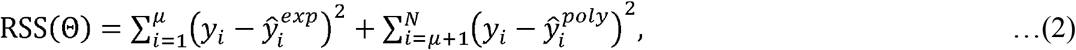

where, 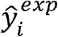 and 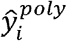 are the estimates value of *y*_*i*_ from the exponential and polynomial regression models, respectively and is the size of the data set.

The estimates of, Θ = {*θ* _1_, *θ* _2_,*µ, β* _0_, *β* _1_, *β* _2_, *β* _3_,……, *β*_*p*_}can be obtained by using the OLS method. We suggest to use the following algorithm while *µ* is kept fixed.

#### Algorithm 1

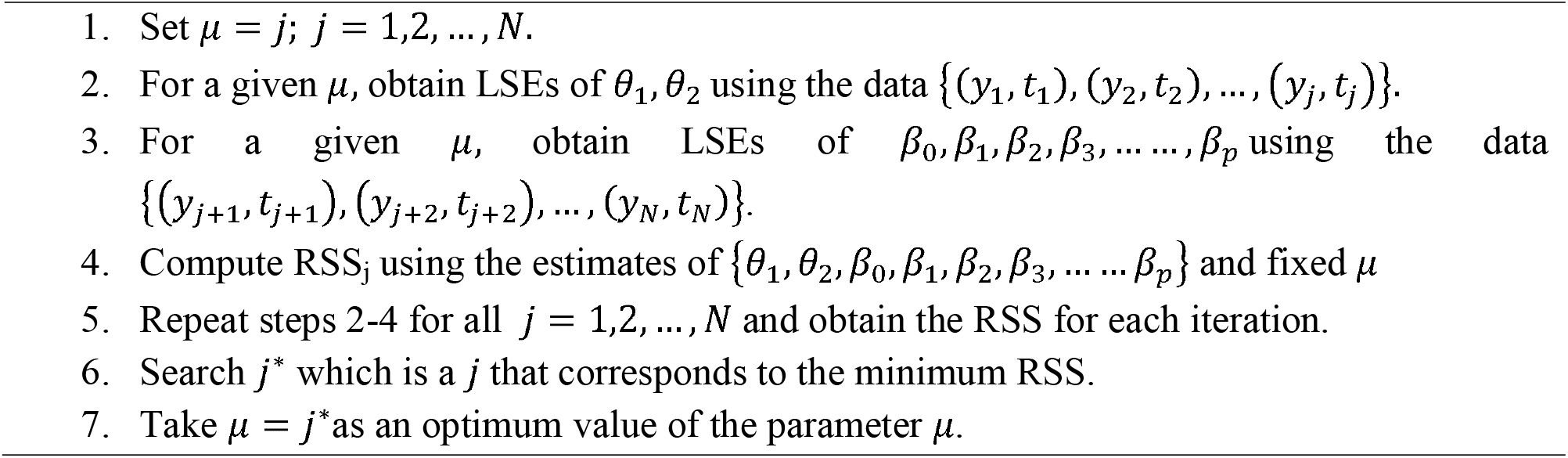

In order to find the optimal value of µ, i.e. the turning point between the exponential and polynomial growth, we will use the technique of minimizing the residual sum squares in section 3.

We will use MAPE (Mean Absolute Percentage Error) in order to evaluate the performance of the model, as given by]

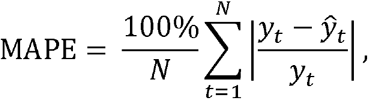

where, *y*_*t*_ is the observed value at time point t and ŷ_*t*_ is an estimate of *y*_*t*_.

In order to make the results easy to interpret, we will also use Accuracy (%) measure defined by

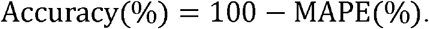

### 2.2 ARIMA Model

The Auto-Regressive Integrated Moving Averages method gauges the strength of one dependent variable relative to other changing variables. It is one of the most used time-series models in diverse fields of data analysis as it takes into account the changing of trends, periodic changes as well as random disturbances in the time-series data. It is used for both better understanding of the data as well as forecasting, see Brockwell et al. [25].

Autoregressive models (AR) is effectively merged with the Moving Averages models (MA) to formulate a useful time-series model, ARIMA model. The *Autoregression (AR)* element of the model shows a changing variable that regresses on its own prior values and the *Moving Average (MA)* element incorporates the dependency between an observation and a residual error from a moving average model applied to prior observations. However, this model can only be applied to stationary data. Since many real-life datasets consist of an element of non-stationarity, in order to model such datasets, ARIMA model was developed. This model is open for non-stationary data as the *Integrated (I)* factor of the model represents the differencing of raw observations to allow the time-series to become stationary.

Here, we may refer the reader to follow Box et al. [26] and Box et al. [27] for more details on ARIMA model, estimation and its application.

The general forms of *AR*(*p*)and *MA* (*q*) models can be respectively represented as the following equations:

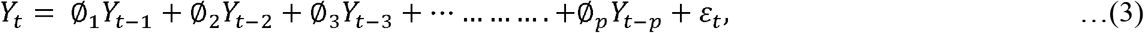

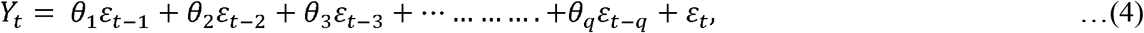

where Øs and θs are autoregressive and moving averages parameters, respectively, *Y*_*t*_ represents value of time-series at time point *t, ε*_*t*_ represents the random disturbance at time point t and is assumed to be independently and identically distributed (i.i.d.) with mean 0 and variance *σ* ^2^.

The *ARMA* (*p,q*) model can be represented as-

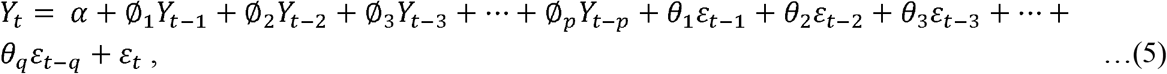

where α is an intercept.

The differenced stationary time-series can be modelled as an ARMA model in order to use ARIMA model on the time-series data, see Ceylan [14], He & Tao [28] and Manikandan et al. [29]. The ARIMA model is generally denoted as *ARIMA* (*p,d,q*), where *p* is the order of auto-regression, d is the degree of difference and q is the order of moving average.

The first step to model the time-series by ARIMA is to determine the time-series data for stationarity. The Augmented Dickey-Fuller (ADF) Test may be applied to determine if the time series after differencing is stationary or not. The ADF test is applied to test the null hypothesis for the presence of a unit root (which indicates non-stationarity of the series).

In order to deduce the *ARIMA* (*p,d,q*) model, we can proceed as follows:

We have the *ARMA* (p’,q) represented as follows (as per Eqn. 5)

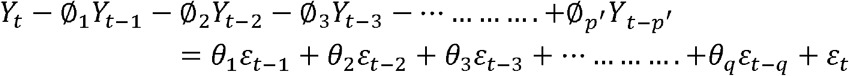

It can be equivalently written as-

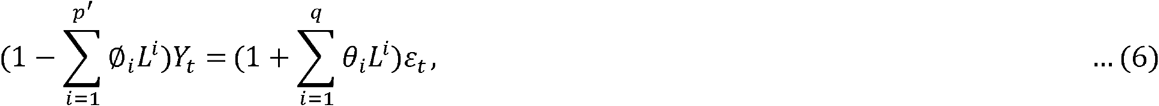

where L is the lag operator, such that-L^a^(*Y*_t_) =*Y*_*t*-a_. Now, assume that the polynomial (1 −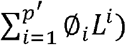 has a unit root (i.e. a factor of (1 − *L*)of multiplicity d. Then, Eqn. (6) can be re-written as-

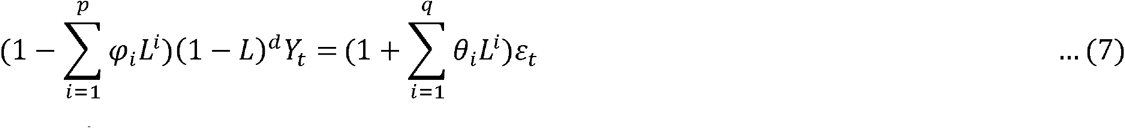

where,*p* ′= *p* −*d*.

This can be generalized as-

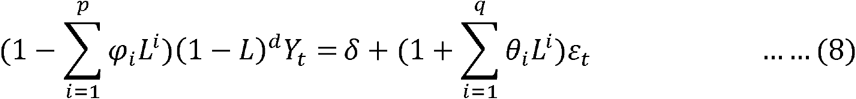

This defines an *ARIMA* (*p,d,q*) process with drift 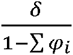.

The second step is to plot the graphs of the Autocorrelation function (ACF) and the Partial Autocorrelation Function (PACF) to determine the most-likely values of *p* and *q*.

The final step is to obtain the optimal values of *p, d* and *q* by using the AIC (Akaike Information Criterion), for more details see https://en.wikipedia.org/wiki/Akaike_information_criterion. These information criteria may be used for selecting the best fitted models. Lower the values of criteria, higher will be its relative quality. The AIC is given by-

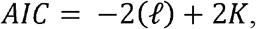

*where K=number of model parameters, ℓ = maximized value of log* − *likeli hood function*.

### 2.3 Exponential Smoothing

Exponential smoothing is one of the simple techniques to model time-series data where the past observations are assigned weights that are exponentially decreasing over time. We propose the following models, for modelling of Covid-19 cases (see Holt [30] and Winters [31]).

For single exponential smoothing, let the raw observations be denoted by {y_*t*_} and {s_*t*_}denote the best estimate of trend at time t. Then, *s*_0_= *y*_0_, *s*_*t*_= *αy*_*t*_+(1− *α*)(*s*_t−1_) where *α ε* (0,1) denotes the data smoothing factor.

For double exponential (Holt-Winters) smoothing, let the raw observations be denoted by {*y_t_*}, smoothened values by {*s_t_*}, and {*b_t_*} denotes the best estimate of trend at time t. Then,

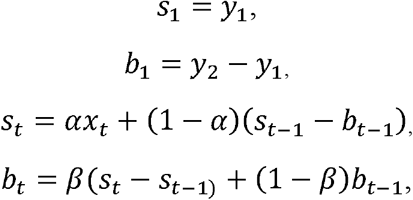

where *α ∈* (0,1) denotes the data smoothing factor and *β ∈* (0,1) denotes the trend smoothing factor. For the forecast at *t* =(*N* + *m*) days, *F*_*N+m*_ is calculated by

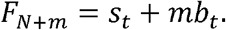

## 3. Analysis of Covid-19 cases in India

For this study, we have used the data available at GitHub, provided by Centre for Systems Science and Engineering (CSSE) at John Hopkins University (see [32]). For this study, we use R software. (see R Core Team [33]).

### 3.1 Exponential-Polynomial Regressions

We know that at the beginning of the spread of the disease in India, the growth was exponential and after some time, it was shifted to polynomial. We first obtain optimum turning point of the growth, i.e. when did the growth rate of the disease shifted to polynomial regime from the exponential. We consider both quadratic and cubic regression model for second part of the data. We will also discuss the types of polynomial growth (with their equations) in India.

In order to find the turning point of the growth curve, we follow the Algorithm 1, given in the previous section. Using that, we evaluate the RSS for all the days (from March 4^th^) and find the date on which it is minimum. The change points of growth curve for cubic and quadratic regressions are presented in Figure 1 depending upon the size of the data set. ***From Figure 1, we can confirm that the growth rate of Covid-19 cases was exponential till April 5***^***th***^ ***and then after it follows the polynomial growth regime while we use the Covid-19 cases till May 2***^***nd***^.

**Figure 1:**
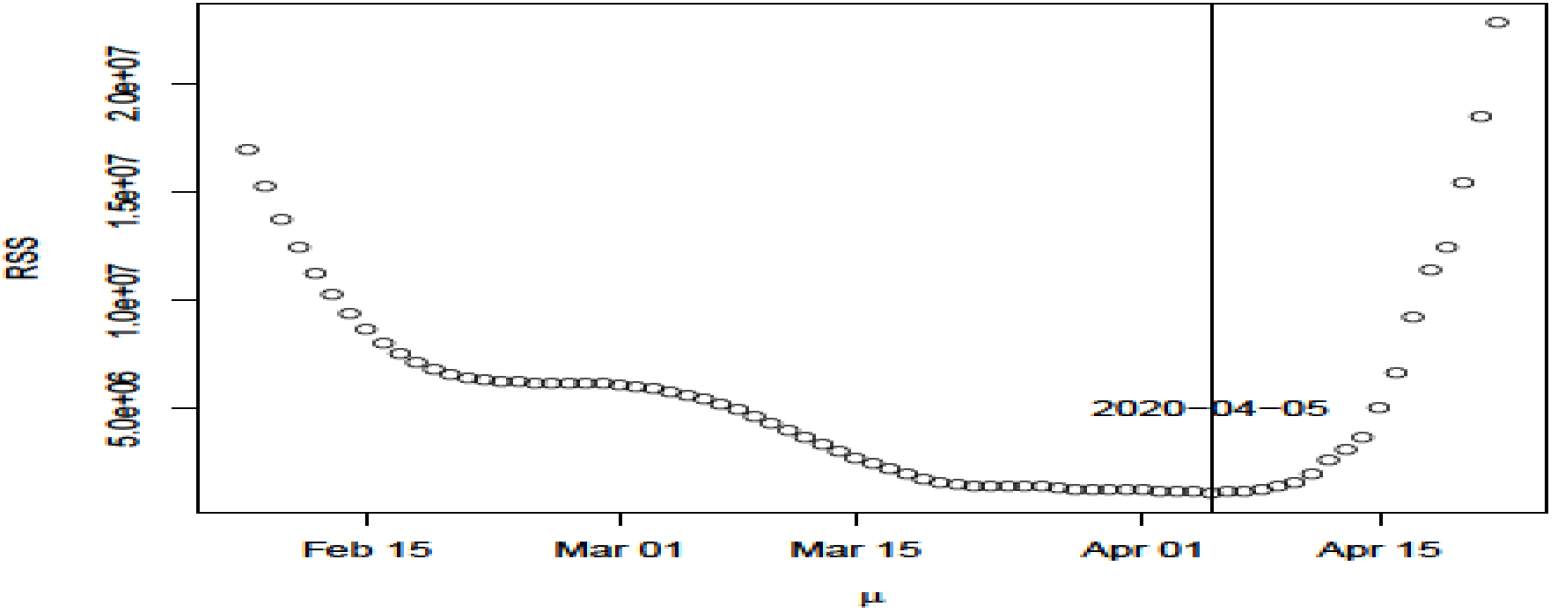
Trend of RSS and optimum µ for exponential-quadratic regression model.

We call the region of exponential growth in India as Region I. The coefficients of the model are presented in Table 2.

**Table 1:** Turning point of growth curve for cubic and quadratic regression beyond change point using the Covid-19 cases from 4^th^ March to a given day.

| Day | Change Point |  |
| --- | --- | --- |
|  | Cubic | Quadratic |
| 25<sup>th</sup> April | 5<sup>th</sup> April | 5<sup>th</sup> April |
| 30<sup>th</sup> April | 5<sup>th</sup> April | 5<sup>th</sup> April |
| 2<sup>rd</sup> May | 5<sup>th</sup> April | 5<sup>th</sup> April |
| 3<sup>rd</sup> May | 7<sup>th</sup> April | 5<sup>th</sup> April |
| 5<sup>th</sup> May | 10<sup>th</sup> April | 11<sup>th</sup> April |
| 10<sup>th</sup> May | 10<sup>th</sup> April | 18<sup>th</sup> April |

**Table 2:** Regression Table for Region I (Exponential Regression)

| Parameter | Coefficients | S.E. | t | PV |
| --- | --- | --- | --- | --- |
| $\theta_1$ | 16.543 | 1.969 | 8.40 | 1.7e-09 |
| $\theta_2$ | 0.163 | 0.00389 | 41.97 | 2e-16 |

We see that after the exponential regime (till April 5^th^), the growth curve follows a polynomial growth till May 2^nd^. After this, we again see a change in the behavior of the growth curve. In Tables 3, 4, 5 and 6, we try to model these growth curves through regression analysis.

**Table 3:** Regression models fitting for Region II (5^th^ April – 2^nd^ May).

| Model | Parameter | OLS Estimates | S.E. | t | PV | R<sup>2</sup> | (F statistic, PV) |
| --- | --- | --- | --- | --- | --- | --- | --- |
| Linear | $\beta_0$ | -43427.02 | 1889.22 | -22.99 | <2e-16 | 0.9773 | (1119, <2e-16) |
| | $\beta_1$ | 1326.49 | 39.66 | 3.45 | <2e-16 | | |
| Quadratic | $\beta_0$ | 17410.66 | 1501.022 | 11.60 | 2.52e-11 | 0.9997 | (3.901e+04, <2.26e-16) |
| | $\beta_1$ | -1335.45 | 65.087 | -20.52 | <2e-16 | | |
| | $\beta_2$ | 28.32 | 0.6905 | 41.01 | <2e-16 | | |
| Cubic | $\beta_0$ | 6196.57 | 10073.87 | 0.615 | 0.545 | 0.9997 | (2.63e+04, <2e-16) |
| | $\beta_1$ | -594.89 | 661.096 | -0.9 | 0.378 | | |
| | $\beta_2$ | 12.29 | 14.2489 | 0.863 | 0.397 | | |
| | $\beta_3$ | 0.113 | 0.1009 | 1.126 | 0.272 | | |

**Table 4:** Regression models fitting Table for Region III (3^rd^ May – 15^th^ May).

| Model | Parameter | OLS Estimates | S.E. | t | PV | R<sup>2</sup> | (F statistic, PV) |
| --- | --- | --- | --- | --- | --- | --- | --- |
| Linear | $\beta_0$ | -2998284.88 | 3244.99 | -91.62 | <2e-16 | 0.9990 | (1.264e+04, <2e-16) |
| | $\beta_1$ | 3584.12 | 32.11 | 111.63 | <2e-16 | | |
| Quadratic | $\beta_0$ | -23424.15 | 56089.18 | -0.418 | 0.68505 | 0.9997 | (1.329e+04, <2.26e-16) |
| | $\beta_1$ | -1866.15 | 1111.75 | -1.679 | 0.12416 | | |
| | $\beta_2$ | 26.98 | 5.50 | 4.903 | 0.00062 | | |
| Cubic | $\beta_0$ | -8.967e+05 | 1.837e+06 | -0.488 | 0.637 | 0.9997 | (1.187e+04, <2e-16) |
| | $\beta_1$ | 2.411e+04 | 5.464e+04 | 0.441 | 0.669 | | |
| | $\beta_2$ | -2.305e+02 | 5.413e+02 | -0.426 | 0.680 | | |
| | $\beta_3$ | 8.497e-01 | 1.786e+00 | 0.476 | 0.646 | | |

**Table 5:** Regression models fitting Table for Region IV (16^th^ May-31^st^ May).

| Model | Parameter | OLS Estimates | S.E. | t | PV | R<sup>2</sup> | (F statistic, PV) |
| --- | --- | --- | --- | --- | --- | --- | --- |
| Linear | $\beta_0$ | -403709.8 | 9765.5 | -41.34 | 4.92e-16 | 0.9951 | (3069, <2.2e-16) |
| | $\beta_1$ | 6627.8 | 119.6 | 55.40 | <2e-16 | | |
| Quadratic | $\beta_0$ | 296345.02 | 50252.76 | 5.897 | 5.26e-05 | 0.9997 | (2.285e+04, <2.26e-16) |
| | $\beta_1$ | -10606.59 | 1235.97 | -8.582 | 1.03e-06 | | |
| | $\beta_2$ | 105.73 | 7.58 | 13.948 | 3.37e-09 | | |
| Cubic | $\beta_0$ | -7.339+05 | 1.025e+06 | -0.716 | 0.488 | 0.9997 | (1.525e+04, <2e-16) |
| | $\beta_1$ | 2.746e+04 | 3.783e+04 | 0.726 | 0.482 | | |
| | $\beta_2$ | -3.623e+02 | 4.649e+02 | -0.779 | 0.451 | | |
| | $\beta_3$ | 1.914e+00 | 1.901e+00 | 1.007 | 0.334 | | |

**Table 6:**
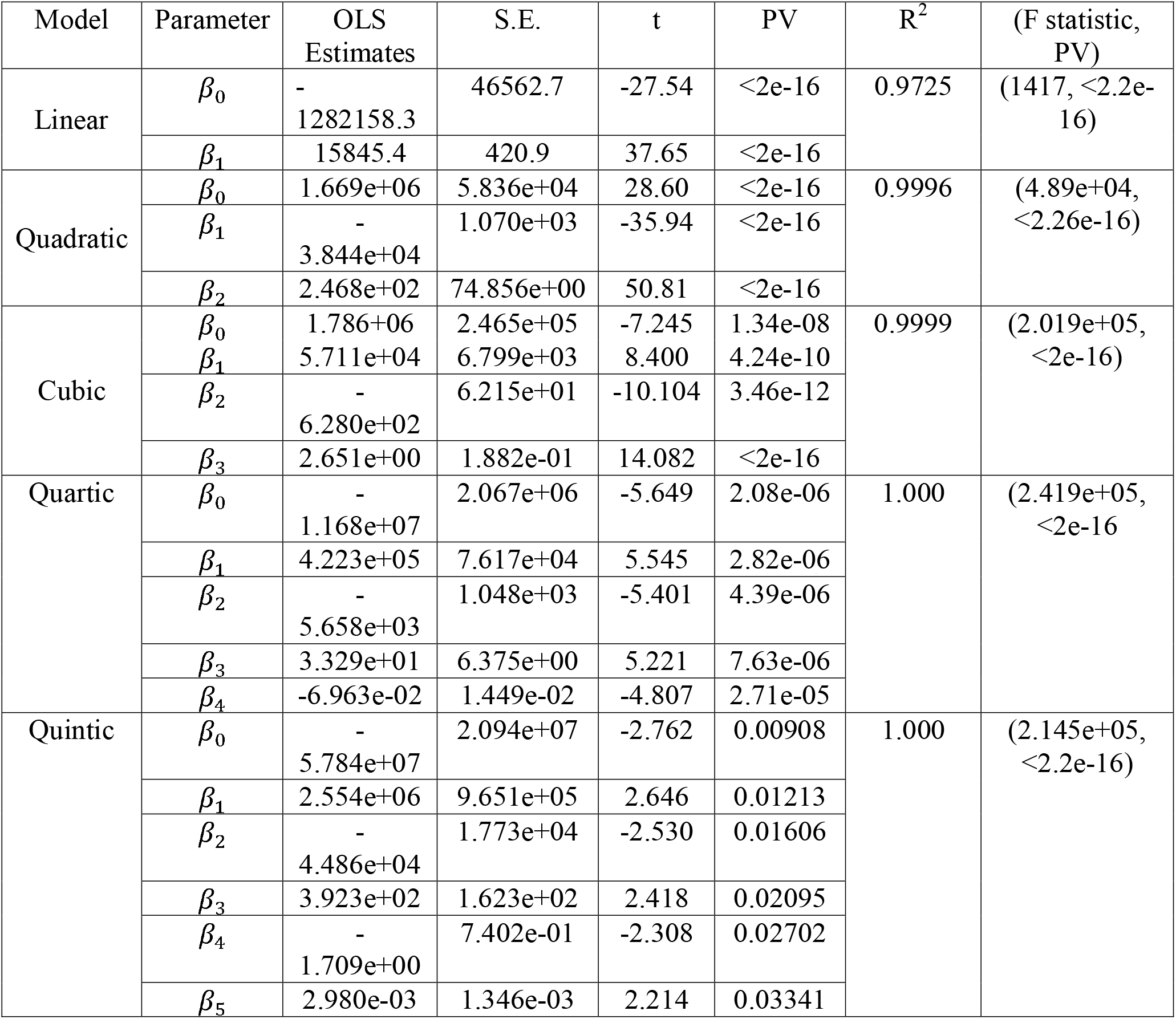
Regression models fitting Table for Region V (1^st^ June-11^th^ July).

| Model | Parameter | OLS Estimates | S.E. | t | PV | R<sup>2</sup> | (F statistic, PV) |
| --- | --- | --- | --- | --- | --- | --- | --- |
| Linear | $\beta_0$ | -1282158.3 | 46562.7 | -27.54 | <2e-16 | 0.9725 | (1417, <2.2e-16) |
| | $\beta_1$ | 15845.4 | 420.9 | 37.65 | <2e-16 | | |
| Quadratic | $\beta_0$ | 1.669e+06 | 5.836e+04 | 28.60 | <2e-16 | 0.9996 | (4.89e+04, <2.26e-16) |
| | $\beta_1$ | -3.844e+04 | 1.070e+03 | -35.94 | <2e-16 | | |
| | $\beta_2$ | 2.468e+02 | 74.856e+00 | 50.81 | <2e-16 | | |
| Cubic | $\beta_0$ | 1.786e+06 | 2.465e+05 | -7.245 | 1.34e-08 | 0.9999 | (2.019e+05, <2e-16) |
| | $\beta_1$ | 5.711e+04 | 6.799e+03 | 8.400 | 4.24e-10 | | |
| | $\beta_2$ | -6.280e+02 | 6.215e+01 | -10.104 | 3.46e-12 | | |
| | $\beta_3$ | 2.651e+00 | 1.882e-01 | 14.082 | <2e-16 | | |
| Quartic | $\beta_0$ | -1.168e+07 | 2.067e+06 | -5.649 | 2.08e-06 | 1.000 | (2.419e+05, <2e-16) |
| | $\beta_1$ | 4.223e+05 | 7.617e+04 | 5.545 | 2.82e-06 | | |
| | $\beta_2$ | -5.658e+03 | 1.048e+03 | -5.401 | 4.39e-06 | | |
| | $\beta_3$ | 3.329e+01 | 6.375e+00 | 5.221 | 7.63e-06 | | |
| | $\beta_4$ | -6.963e-02 | 1.449e-02 | -4.807 | 2.71e-05 | | |
| Quintic | $\beta_0$ | -5.784e+07 | 2.094e+07 | -2.762 | 0.00908 | 1.000 | (2.145e+05, <2.2e-16) |
| | $\beta_1$ | 2.554e+06 | 9.651e+05 | 2.646 | 0.01213 | | |
| | $\beta_2$ | -4.486e+04 | 1.773e+04 | -2.530 | 0.01606 | | |
| | $\beta_3$ | 3.923e+02 | 1.623e+02 | 2.418 | 0.02095 | | |
| | $\beta_4$ | -1.709e+00 | 7.402e-01 | -2.308 | 0.02702 | | |
| | $\beta_5$ | 2.980e-03 | 1.346e-03 | 2.214 | 0.03341 | | |

Having evaluated the coefficients for various models (i.e. linear, quadratic and cubic) as well as the important statistics (i.e. R^2^ values, p-values of the models as well as individual coefficients and F-statistic), we will select the best fitting models. In order to select the best fitting models for Region II (April 6^th^ to May 2^nd^); III (May 3^rd^ to May 15^th^), IV (May 16^th^ to May 31^st^) and V (June 1^st^ to July 11^th^), we have the following steps. We select that model which has *high R*^*2*^ *values, significant p-value, high F-statistic* and where the *p-values of all the variables are significant*.

We see for **Region II**, from Table 3 that the linear model is having a relatively lower F-statistic and R^2^ values in comparison to the Quadratic and Cubic models. So, we eliminate the possibility of linear fitting. Further, we see that the p-values, F-statistics and the R^2^ values are quite significant in both Quadratic as well as the Cubic models. But, if we look at the individual p-values of the coefficients, we see that the individual p-values are not significant for the Cubic model. On the other hand, the individual p-values are significant for the Quadratic model. Thus, we can conclude that the *Quadratic* model is the best fitting model for Region II (April 6^th^ to May 2^nd^).

For **Region III**, from Table 4, that all the three models have high F-statistic values, high p-values and high R^2^ values. But we notice that the coefficient individual p-values are not significant in both Quadratic and Cubic models. Thus, we conclude that the *Linear* model is the best fitting model for Region III (May 3^rd^ to May 15^th^).

For **Region IV**, from Table 5, we see that the R^2^ values for all the models are very high. All the models also have significant p-values. The F-statistic of both Quadratic and Cubic models are also high. But, the coefficient individual p-values are not significant in the Cubic model. Thus, we conclude that the *Quadratic* model is the best fitting model for Region IV (May 16^th^ to May 31^st^).

For **Region V**, from Table 6, we see that the R^2^ values of all the models are very high (quadratic, cubic, quartic and quintic models have exceptionally high). All the models also have significant p-values. The F-statistic of Quadratic, Cubic, Quartic and Quintic models is high. F-statistic value of Quartic model is the highest. The coefficient individual p-values of Quartic model are also significant. Thus, we conclude that the *Quartic* model is the best fitting model for Region V (June 1^st^ to July 11^th^)

**Note:** For Region V, due to spike in the cases, we also checked the fitting of exponential regression in this region, see Table 7. The RSE for Exponential Regression is 4178 and MAPE (%) is 0.85%. Both of these values are quite larger than those of Quartic model. (Refer Table 9 for RSE and MAPE values of Quartic model in Region V). Thus, we conclude that Quartic model is the best-fitting model for Region V (1^st^ June to 11^th^ June).

**Table 7:** Parameters for Exponential Regression in Region V

| Parameter | OLS Estimate | S.E. | T | p-value |
| --- | --- | --- | --- | --- |
| $\theta_1$ | 8.718e+03 | 1.385e+02 | 62.95 | <2e-16 |
| $\theta_2$ | 3.530e-02 | 1.333e-04 | 264.69 | <2e-16 |

All the ANOVA (Table 8) for Region II, III, IV and V suggest significant p-values for its coefficients and suggest that the models fit well the respective regions.

**Table 8:**
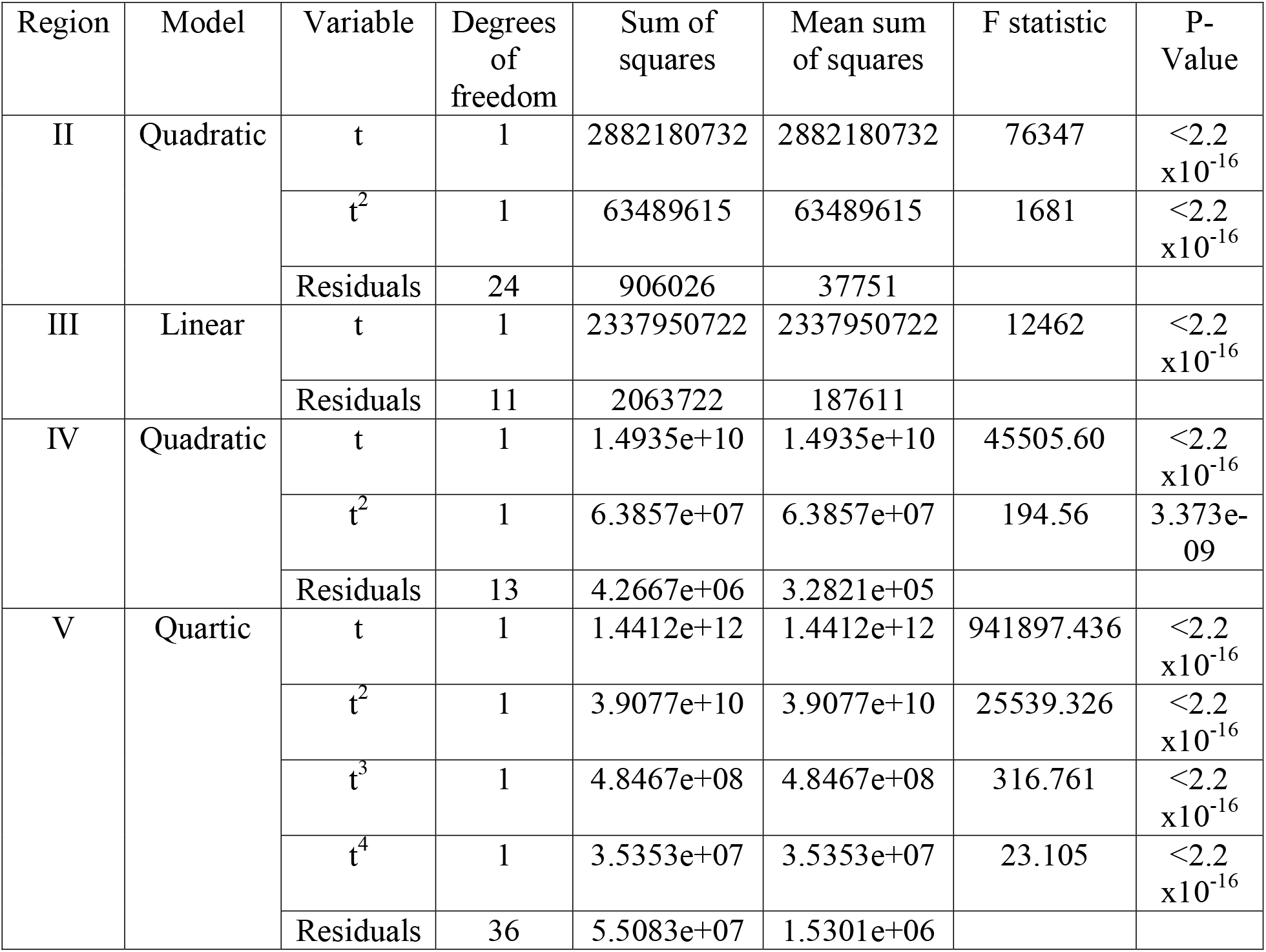
ANOVA Table for Region II (Quadratic Regression), III (Linear Regression), IV (Quadratic Regression) and V (Quantic Regression)

| Region | Model | Variable | Degrees of freedom | Sum of squares | Mean sum of squares | F statistic | P-Value |
| --- | --- | --- | --- | --- | --- | --- | --- |
| II | Quadratic | t | 1 | 2882180732 | 2882180732 | 76347 | $<2.2 \times 10^{-16}$ |
| | | $t^2$ | 1 | 63489615 | 63489615 | 1681 | $<2.2 \times 10^{-16}$ |
|  |  | Residuals | 24 | 906026 | 37751 |  |  |
| III | Linear | t | 1 | 2337950722 | 2337950722 | 12462 | $<2.2 \times 10^{-16}$ |
|  |  | Residuals | 11 | 2063722 | 187611 |  |  |
| IV | Quadratic | t | 1 | 1.4935e+10 | 1.4935e+10 | 45505.60 | $<2.2 \times 10^{-16}$ |
| | | $t^2$ | 1 | 6.3857e+07 | 6.3857e+07 | 194.56 | 3.373e-09 |
|  |  | Residuals | 13 | 4.2667e+06 | 3.2821e+05 |  |  |
| V | Quartic | t | 1 | 1.4412e+12 | 1.4412e+12 | 941897.436 | $<2.2 \times 10^{-16}$ |
| | | $t^2$ | 1 | 3.9077e+10 | 3.9077e+10 | 25539.326 | $<2.2 \times 10^{-16}$ |
| | | $t^3$ | 1 | 4.8467e+08 | 4.8467e+08 | 316.761 | $<2.2 \times 10^{-16}$ |
| | | $t^4$ | 1 | 3.5353e+07 | 3.5353e+07 | 23.105 | $<2.2 \times 10^{-16}$ |
|  |  | Residuals | 36 | 5.5083e+07 | 1.5301e+06 |  |  |

**Table 9:** Course of Covid-19 growth in India (March 4^th^ to July 11^th^)

| Region | Dates | Best fitted model | MAPE (%) | RSE |
| --- | --- | --- | --- | --- |
| I | March 4<sup>th</sup> to April 5<sup>th</sup> | $y(t) = 16.54 \times e^{0.163t}$ | 8.60 | 81.66 |
| II | April 6<sup>th</sup> to May 2<sup>nd</sup> | $y(t) = 17410.67 - 1335.45t + 28.32t^2$ | 0.83 | 194.3 |
| III | May 3<sup>rd</sup> to May 15<sup>th</sup> | $y(t) = -29825 + 3584t$ | 0.49 | 433.1 |
| IV | May 16<sup>th</sup> to May 31<sup>st</sup> | $y(t) = 296345.02 - 10606.59t + 105.73t^2$ | 0.31 | 572.6 |
| V | June 1<sup>st</sup> to July 11<sup>th</sup> | $y(t) = -1.168 \times 10^7 + 4.223 \times 10^5t - 5.658 \times 10^3t^2 + 33.29t^3 - 0.06963t^4$ | 0.21 | 1237 |

*Thus, according to our study, the growth of the virus was exponentially increasing from March 4*^*th*^ *to April 5*^*th*^. *Then after, the covid 19 cases grew by following a quadratic rate from April 6*^*th*^ *to May 2*^*nd*^. *After May 3*^*rd*^, *we experienced a linear growth. But after May 15*^*th*^ *to May 31*^*st*^, *we experienced a sudden rise in the rate of growth of the pandemic and have seen quadratic growth again. Further, for the period of June 1*^*st*^ *to July 11*^*th*^, *we see experienced a quartic (4-degree polynomial) growth, which is very alarming. (See Table 9 for best fitted regression models)*. Figure 2 shows the best fitted regression models to the daily cumulative cases of Covid-19 in India from March 4^th^ to July 11^th^.

**Figure 2:**
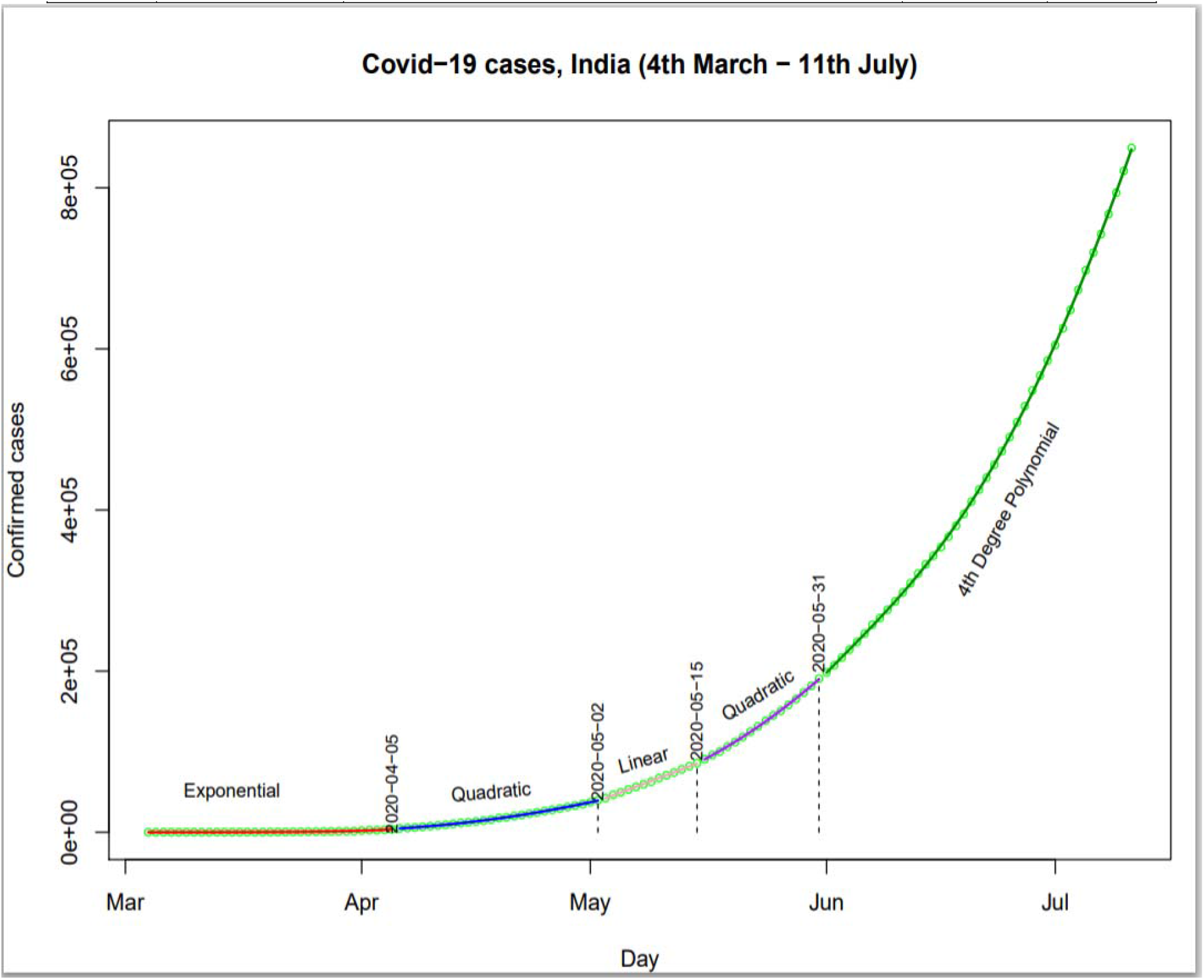
Fitted Regression models (given in Table 9) to the daily cumulative cases of Covid-19 in India till July 11^th^ (circle represents the observed value).

### 3.2 Time series Models fitting

We use cumulative confirmed cases from March 4^th^ to July 10^th^ for fitting time series models and data from July 11^th^ to July 14^th^ is used for model validation.

First, we check the stationarity of the data using the ADF Test. ADF test statistic is 6.3915 with p-value 0.99, which indicates that the growth of Covid-19 cases is not stationary. The ARIMA models may be useful over the ARMA models. In order to make time series stationary, we may apply difference operator. In Figure 6, we can see that 2^nd^ order differencing make the series almost stationary. Thus, we can take *d* =2. The ACF and PACF plots are shown in Figure 3. ACF plot indicates that there is high autocorrelation (more than 0.5) up to time lag 15 days.

**Figure 3:**
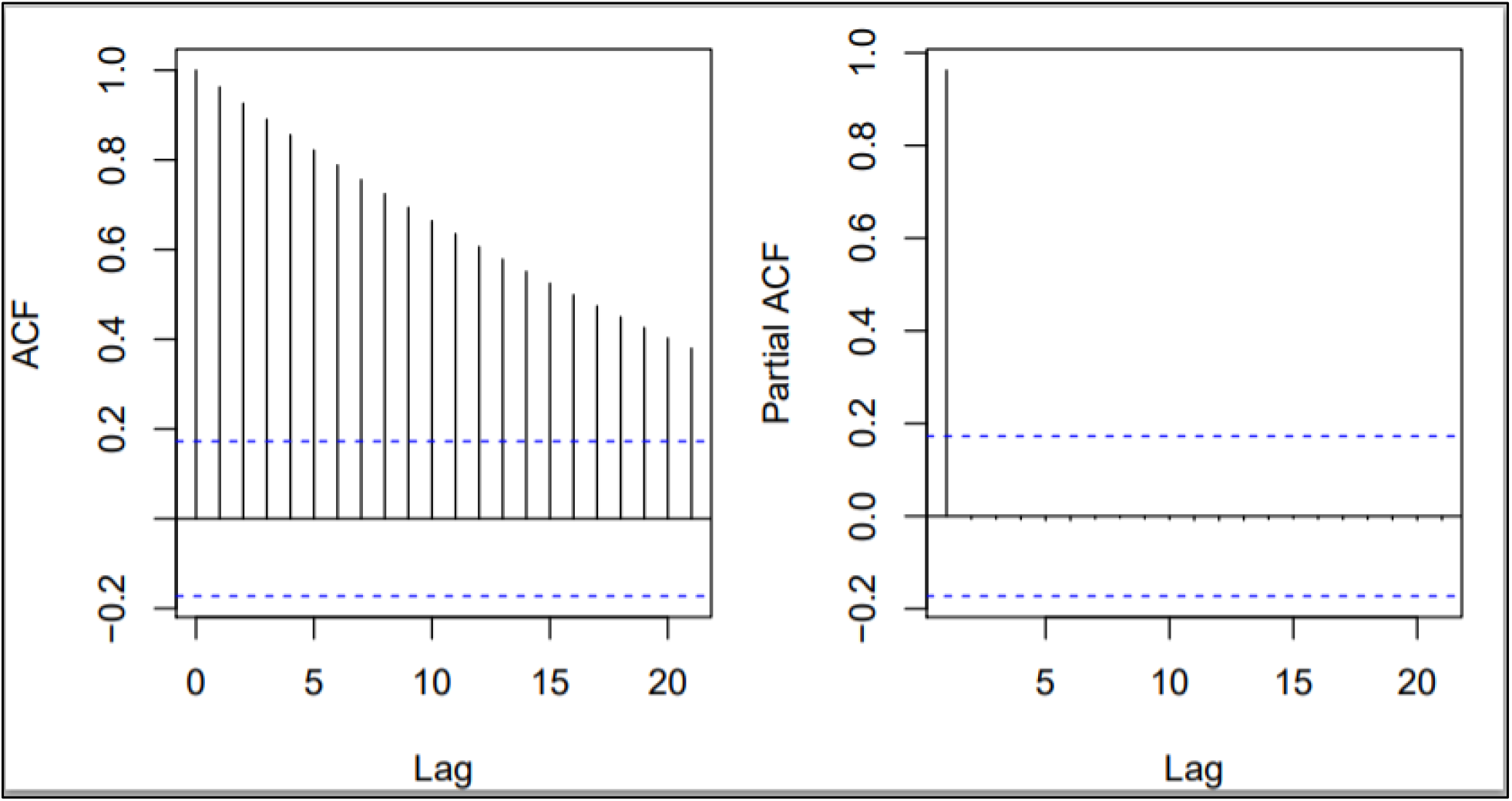
ACF and PACF for Covid-19 cases in India (4^th^ March to 10^th^ July)

We then obtain the optimal ARIMA parameters (*p, d, q*) by using the AIC. As suggested by Box et al. [26, 27], we take various possible combinations of *p* =1,2,…,5, *q* =1,2,…,5 and *d* =1,2 and compute the AIC.

According to the AIC, the ARIMA (5, 2, 5) is the best fitted model for the Covid-19 cases, India (see Table 10). Estimates of ARIMA (5, 2, 5) parameters and MAPE are shown in Table 11.

**Table 10:** AIC for ARIMA models for Covid-19 cases, India (4^th^ March to 10^th^ July)

| Model Parameter |  |  | AIC |
| --- | --- | --- | --- |
| p | d | q |  |
| 1 | 1 | 0 | 2064.921 |
| 2 | 1 | 0 | 2065.752 |
| 3 | 1 | 0 | 2067.442 |
| 2 | 1 | 2 | 2063.402 |
| 3 | 1 | 5 | 2055.702 |
| 2 | 0 | 1 | 2070.555 |
| 1 | 2 | 1 | 2043.075 |
| 2 | 2 | 1 | 2044.97 |
| 1 | 2 | 2 | 2045 |
| 2 | 2 | 2 | 2022.933 |
| 3 | 2 | 2 | 2048.967 |
| 3 | 2 | 3 | 2021.517 |
| **5** | **2** | **5** | **2001.998** |
| 2 | 2 | 5 | 2012.213 |
| 5 | 2 | 4 | 2008.532 |
| 3 | 2 | 5 | 2013.83 |

**Table 11:**
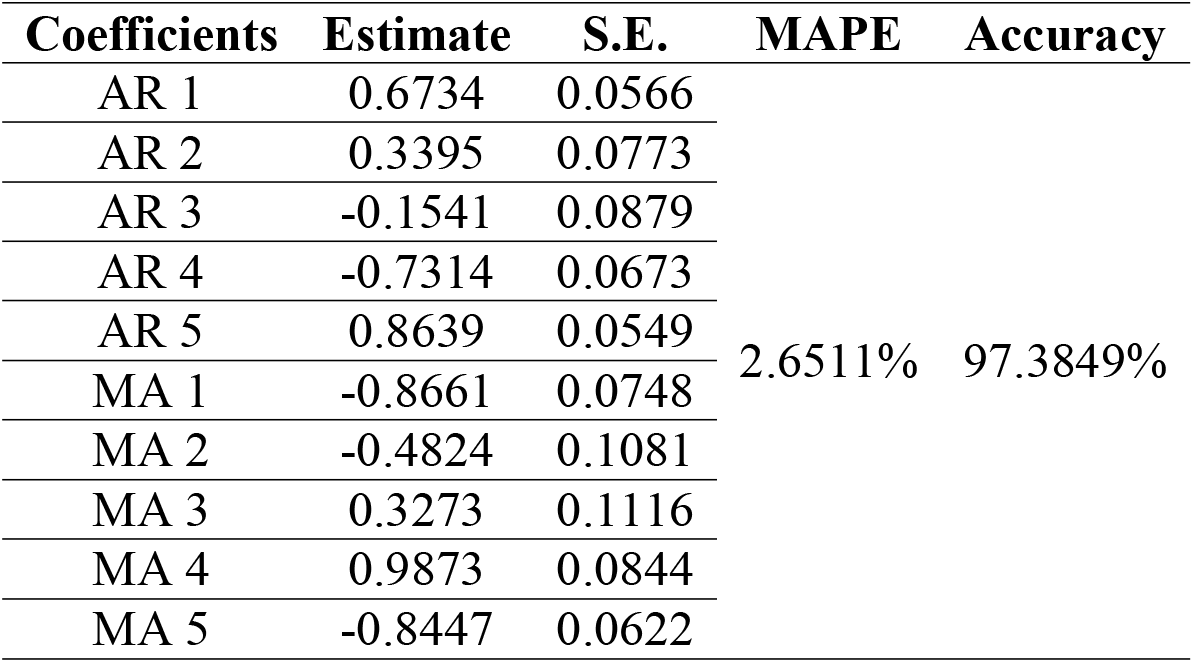
Estimates of ARIMA (5, 2, 5) parameters and MAPE (4^th^ March to 10^th^ July)

| Coefficients | Estimate | S.E. | MAPE | Accuracy |
| --- | --- | --- | --- | --- |
| AR 1 | 0.6734 | 0.0566 | 2.6511% | 97.3849% |
| AR 2 | 0.3395 | 0.0773 |  |  |
| AR 3 | -0.1541 | 0.0879 |  |  |
| AR 4 | -0.7314 | 0.0673 |  |  |
| AR 5 | 0.8639 | 0.0549 |  |  |
| MA 1 | -0.8661 | 0.0748 |  |  |
| MA 2 | -0.4824 | 0.1081 |  |  |
| MA 3 | 0.3273 | 0.1116 |  |  |
| MA 4 | 0.9873 | 0.0844 |  |  |
| MA 5 | -0.8447 | 0.0622 |  |  |

Estimates of the Holt-Winters exponential smoothing and exponential smoothing models are given in Table 12. According to the MAPE and accuracy measures, the ARIMA (5, 2, 5) is a better model than the Holt-Winters exponential smoothing and usual exponential smoothing models. From this, we can conclude that the ARIMA model gives the best fit for the cases of Covid-19, followed by Holt-Winters model. The forecasting values along with 95% confidence intervals are shown in Table 13.

**Table 12:** Estimates and MAPE of exponential smoothing models (4^th^ March to 10^th^ July).

| Model | Parameter | Estimate | MAPE | Accuracy |
| --- | --- | --- | --- | --- |
| Holt-Winters Exponential Smoothing | $\alpha$ | 1 | 2.8911% | 97.1089% |
| | $\beta$ | 1 | | |
|  | a | 820916 |  |  |
|  | b | 27114 |  |  |
| Exponential Smoothing | $\alpha$ | 0.9999 | 7.5384% | 92.4616% |
|  | a | 820914.7 |  |  |

**Table 13:** Forecast using ARIMA and Holt-Winters models for 10 days

| Day | ARIMA |  |  | Holt-Winters |  |  | Actual |
| --- | --- | --- | --- | --- | --- | --- | --- |
|  | Estimate | Lower | Upper | Estimate | Lower | Upper |  |
| 11<sup>th</sup> July | 848374.1 | 847247.1 | 849501.0 | 848030 | 846636.1 | 849423.9 | 849522 |
| 12<sup>th</sup> July | 875792.9 | 873473.0 | 878112.8 | 875144 | 872027.2 | 878260.8 | 878254 |
| 13<sup>th</sup> July | 903010.8 | 899507.1 | 906514.5 | 902258 | 897042.5 | 907473.5 | 906752 |
| 14<sup>th</sup> July | 930330.7 | 925622.6 | 935038.7 | 929372 | 921737.3 | 937006.7 | 936181 |
| 15<sup>th</sup> July | 958494.9 | 952436.5 | 964553.2 | 956486 | 946148.6 | 966823.4 |  |
| 16<sup>th</sup> July | 987619.1 | 979891.2 | 995347.0 | 983600 | 970303.1 | 996896.9 |  |
| 17<sup>th</sup> July | 1017773.7 | 1007937.9 | 1027609.6 | 1010714 | 994221.2 | 1027206.8 |  |
| 18<sup>th</sup> July | 1048569.9 | 1036227.8 | 1060912.1 | 1037828 | 1017919.2 | 1057736.8 |  |
| 19<sup>th</sup> July | 1079470.6 | 1064335.7 | 1094605.5 | 1064942 | 1041410.4 | 1088473.6 |  |
| 20<sup>th</sup> July | 1110527.9 | 1092443.6 | 1128612.2 | 1092056 | 1064705.9 | 1119406.1 |  |

***Note:** Even though the actual cases are covered in the 95% confidence intervals of the ARIMA and Holt- Winters forecasts, they are seen to be nearer to the Upper Limits of the Confidence Intervals and are deviated from the estimates. It might be possible that in future days, the forecasts might underestimate the actual cases. This might be attributed to the changing pattern of the growth of the pandemic in our country as seen in the regression analysis. Thus, we suggest a segment-wise time-series models to forecast the future cases in a more accurate manner.*

We now present ARIMA and Holt-Winters models for 1^st^ June to 10^th^ July, last segment of the study.

We have seen that our time-series data is non-stationary and thus, we select the most optimal values of (*p,d,q*), which has the least AIC. ACF and PACF plots are given in Figure 5. According to AIC, ARIMA(5, 2, 3) is the best-fitting model for the time-series data from June 1^st^ to July 10^th^, with AIC=634.18. Estimates of ARIMA (5, 2, 3) model with the corresponding MAPE and Accuracy are given in Table 14.

**Table 14:** Estimates of ARIMA (5, 2, 3) parameters and MAPE (1^st^ June to 10^th^ July)

| Coefficients | Estimate | S.E. | MAPE | Accuracy |
| --- | --- | --- | --- | --- |
| AR 1 | 1.9985 | 0.2455 | 0.1364 % | 99.8636 % |
| AR 2 | -2.0142 | 0.4505 |  |  |
| AR 3 | 1.2380 | 0.4533 |  |  |
| AR 4 | -0.6108 | 0.3606 |  |  |
| AR 5 | 0.3742 | 0.1895 |  |  |
| MA 1 | -2.2358 | 0.3847 |  |  |
| MA 2 | 2.0805 | 0.7260 |  |  |
| MA 3 | -0.7309 | 0.3853 |  |  |

Estimates of the Holt-Winters exponential smoothing and exponential smoothing models are given in Table 15. According to the MAPE and accuracy measures, the ARIMA (5, 2, 3) is a better model than the Holt-Winters exponential smoothing and usual exponential smoothing models. From this, we can conclude that the ARIMA model also gives the best fit for the cases of Covid-19 from 1^st^ June to 10^th^ July, followed by Holt-Winters model. The forecasting values along with 95% confidence intervals are shown in Table 16 and Figure 4. We observe that the ARIMA model captures the trend well and estimates the cumulative cases properly.

**Table 15:** Estimates and MAPE of exponential smoothing models (June 1^st^ to July 11^th^).

| Model | Parameter | Estimate | MAPE | Accuracy |
| --- | --- | --- | --- | --- |
| Holt-Winters Exponential Smoothing | $\alpha$ | 1 | 0.2172% | 99.7828% |
| | $\beta$ | 1 | | |
|  | a | 820916 |  |  |
|  | b | 271145 |  |  |
| Exponential Smoothing | $\alpha$ | 0.9999 | 3.5757% | 96.4243% |
|  | a | 820914.70 |  |  |

**Table 16:**
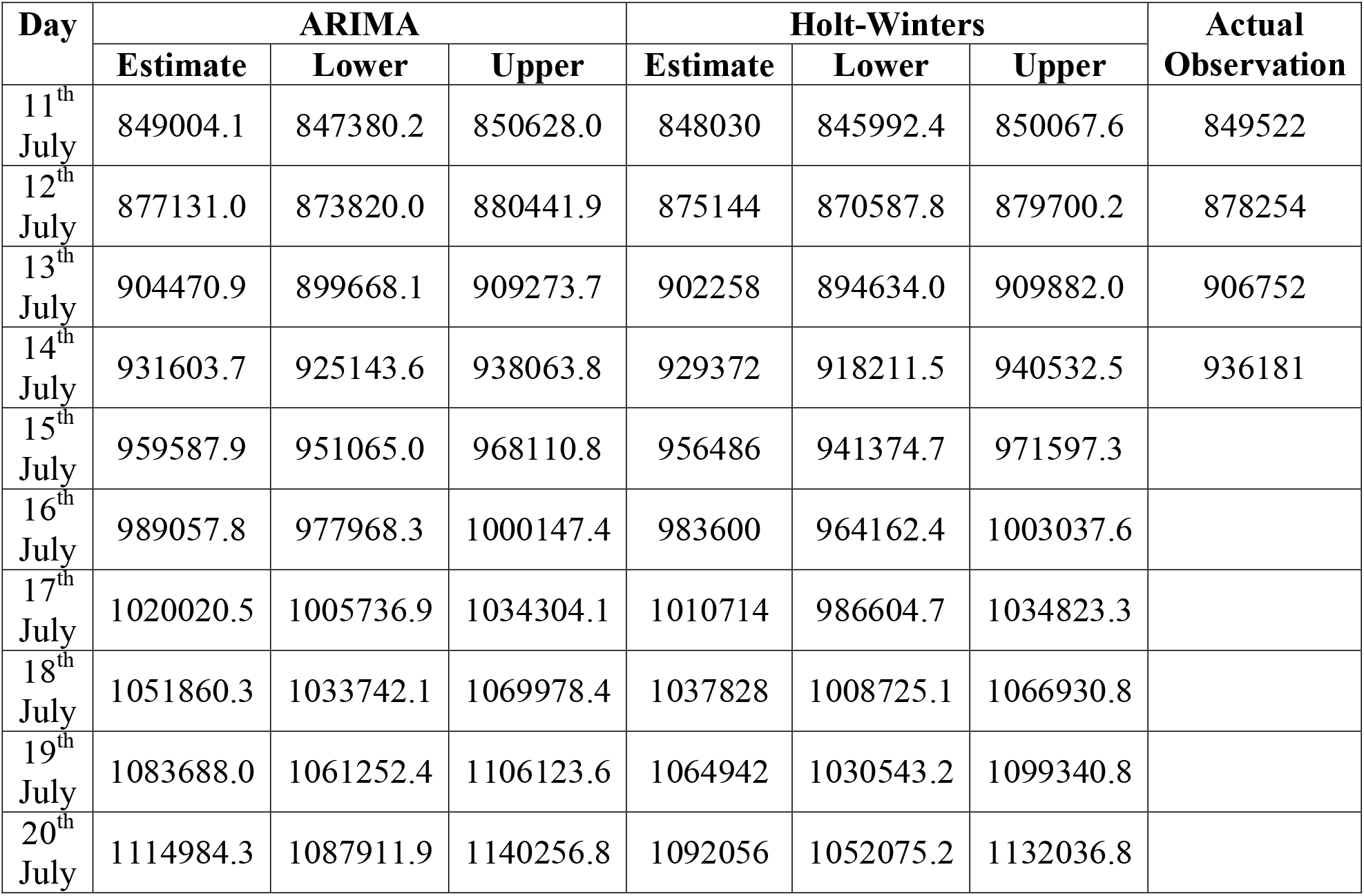
Forecast using ARIMA(5, 2, 3) and Holt-Winters models for 10 days (Model based on data from June 1^st^ to July 11^th^)

**Figure 4.**
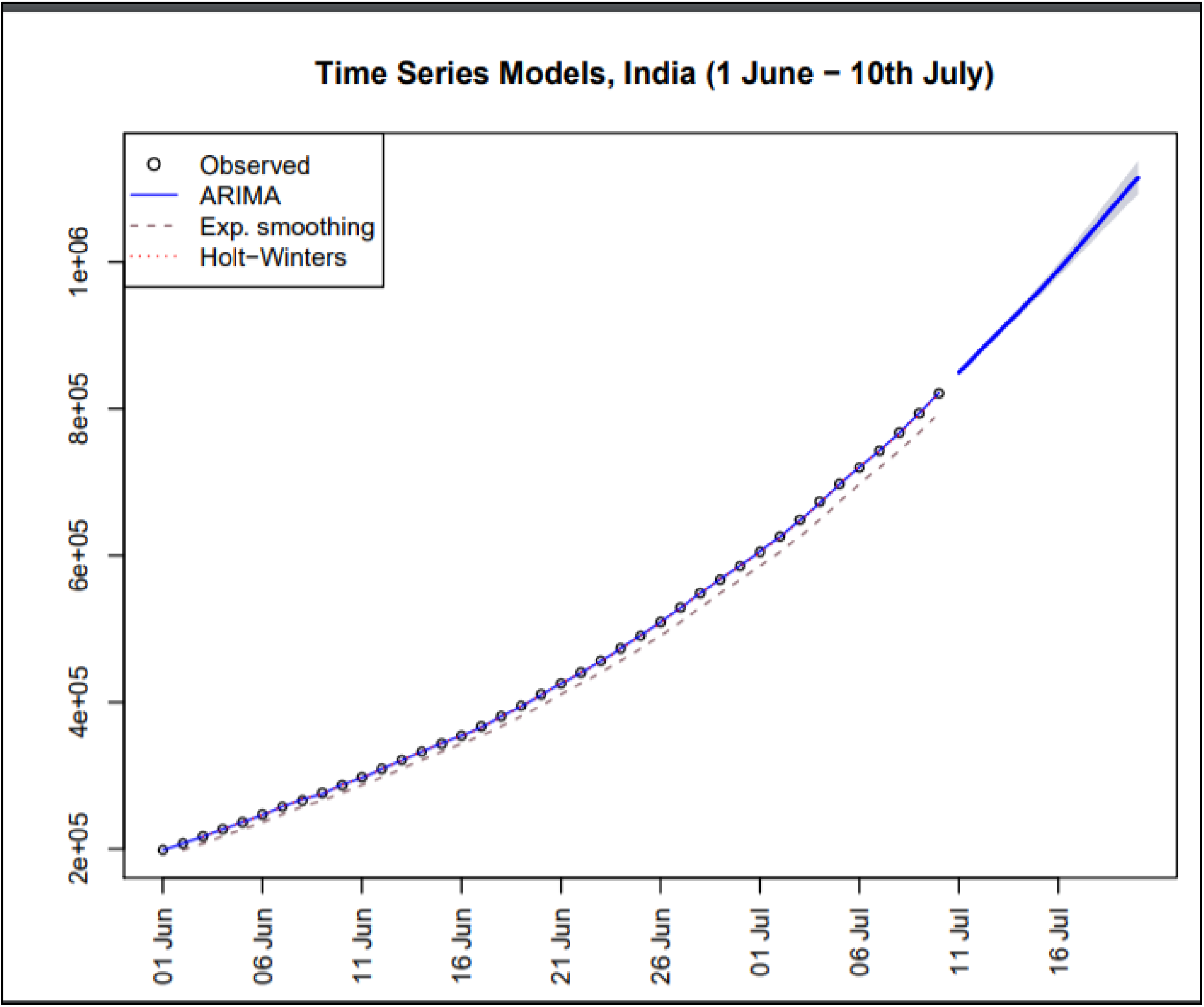
Fitted ARIMA (5, 2, 3) and exponential smoothing models and forecasting from ARIMA for Covid-19 cases in India from June 1^st^ to July 10^th^.

**Figure 5:**
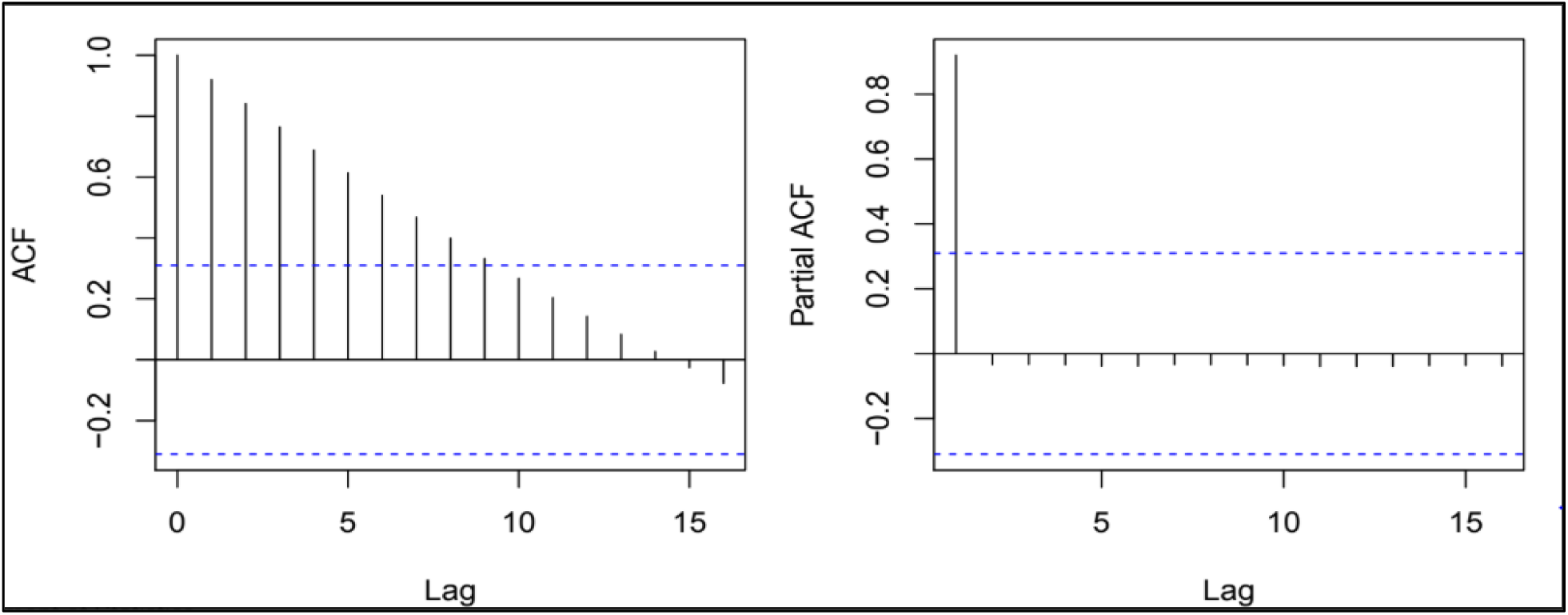
ACF and PACF plots for Covid-19 cases (1^st^ June to 10^th^ July)

**Figure 6:**
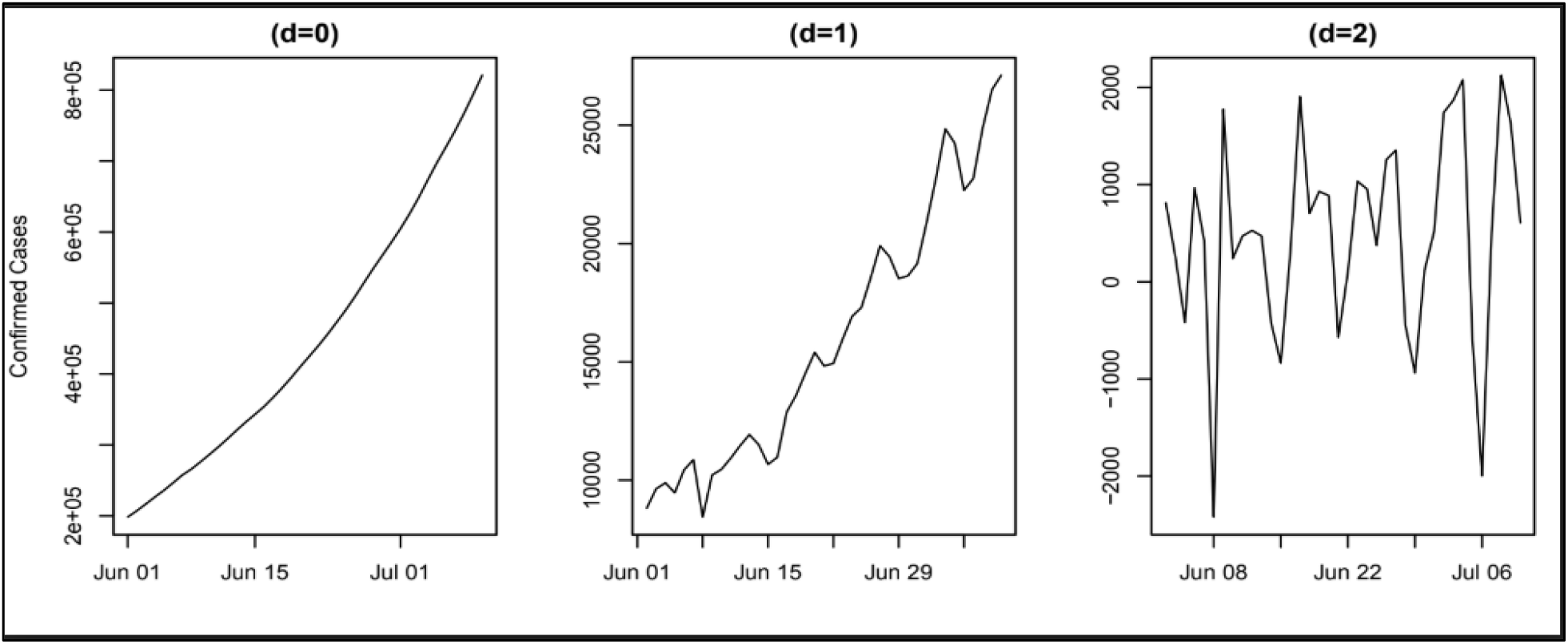
Difference plots for Covid-19 cases (1^st^ June to 10^th^ July)

## 4. Conclusions

From the regression analysis, we conclude that the spread of Covid-19 disease grew exponentially from March 3^rd^ to April 5^th^. Further, from April 6^th^ to May 2^nd^, the cases followed a quadratic regression. From May 3^rd^ to May 15^th^, we see a linear growth of the pandemic with average daily cases of 3584. After May 15^th^ to May 31^st^, we again saw a spike in the cases that lead to a quadratic growth of the pandemic. And, from June 1^st^ to July 11^th^, we saw a major spike in the growth of the pandemic as it has followed quartic growth.

Verma et al. [21] showed the four stages of the epidemic, S1: exponential, S2: power law, S3: linear and S4: flat. We saw that the course of Covid-19 in India followed this regime till May 15^th^. But after the linear trend from May 3^rd^ to May 15^th^, the spread has again reached the quadratic growth and from June 1^st^ to July 11^th^, India is witnessing a quartic growth. This might be attributed to the relaxation of lockdown measures in the country. Though it was much likely that the cases would start to reduce post-linear stage growth as the total cases may start to follow a square root equation, i.e.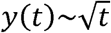. And this might lead to reduction in the daily number of cases (as 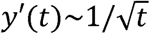,) leading to flattening of the curve. But, due to reduced restrictions, we see a reverse trend, which might be alarming and we suggest the imposition of strict lockdown in order to reverse this trend of pandemic growth. If we continue to open our economy in this way, we might go back to the exponential growth of the pandemic and this would lead to huge destruction to human lives and cause a greater impact on our economy.

We also observe that some cities have been the hotspots of the disease, such as Delhi (more than 1,31,000 cases), Mumbai (more than 94,000 cases), Chennai (more than 78,000 cases), Thane (more than 63,000 cases) etc. as on 14^th^ June, 2020. While the other states and cities have seen a slower growth of the pandemic, these cities have seen explosive growths. Due to the opening of air and rail transport in the country, the virus is likely to spread in the other regions as well as people from these cities (especially metro cities) are travelling to different states. Thus, it is highly advisable that the country should go back to its lockdown phase until we see reduction in trend.

In time series analysis, we conclude that the ARIMA (5, 2, 5) is the best fitting model for the cases of Covid-19 from 4^th^ March to 10^th^ July with an accuracy of 97.38%. The basic exponential smoothing is not very accurate for our case but we see that the Holt-Winters model is around 97.11% accurate. Both ARIMA (5, 2, 5) and Holt-Winters models suggest a rise in the number of cases in the coming days. We observed that both the ARIMA and Holt-Winters models capture the data well and the actual data from 11^th^ July validates the forecasts well as they lie in the predicted confidence intervals. But, while validating the model, the actual values are always near to the Upper Confidence Limits, it might be possible that in further days, our model might underestimate the cases. This might be possible because of the changing trend of the growth of the pandemic in India.

Thus, we used segmented time-series models and took a data from 1^st^ June to 10^th^ July to build separate ARIMA and Holt-Winters models. We concluded that ARIMA (5, 2, 3) is the best fitting model for Covid-19 cases in the given time period with an accuracy of 99.86%. The basic exponential smoothing is not very accurate or this case as well but, the Holt-Winters model is around 99.78% accurate. We also observe that the ARIMA and Holt-Winters models capture the data well and the actual data from 11^th^ July validates the forecasts and lies near to the estimates.

We may also conclude that the cases of Covid-19 will rise in the coming days and the situation may turn alarming if proper measures are not followed. Since the economic activities have started in the country, people need to be more careful while going out. And explosion of the pandemic in the whole country can cause a serious damage to human lives, healthcare system as well as the economy of the country. Thus, there is an urgent need of imposing strict lockdown measures to curb the growth of the pandemic. We must also learn to lead our lives by following all the precautions even if the lockdown restrictions are relaxed and the economic activities are resumed.

## Data Availability

The data is available at GitHub, provided by John Hopkins University. 15.https://github.com/CSSEGISandData/COVID-19/blob/master/csse_covid_19_data/csse_covid_19_time_series/time_series_covid19_confirmed_global.csv

## Acknowledgments

Dr. Vikas Kumar Sharma greatly acknowledges the financial support from Science and Engineering Research Board, Department of Science & Technology, Govt. of India, under the scheme Early Career Research Award (file no.: ECR/2017/002416).

